## Supplementary Information for "Influenza Hospitalisations in England during the 2022/23 Season: do different data sources drive divergence in modelled waves? A comparison of surveillance and administrative data"

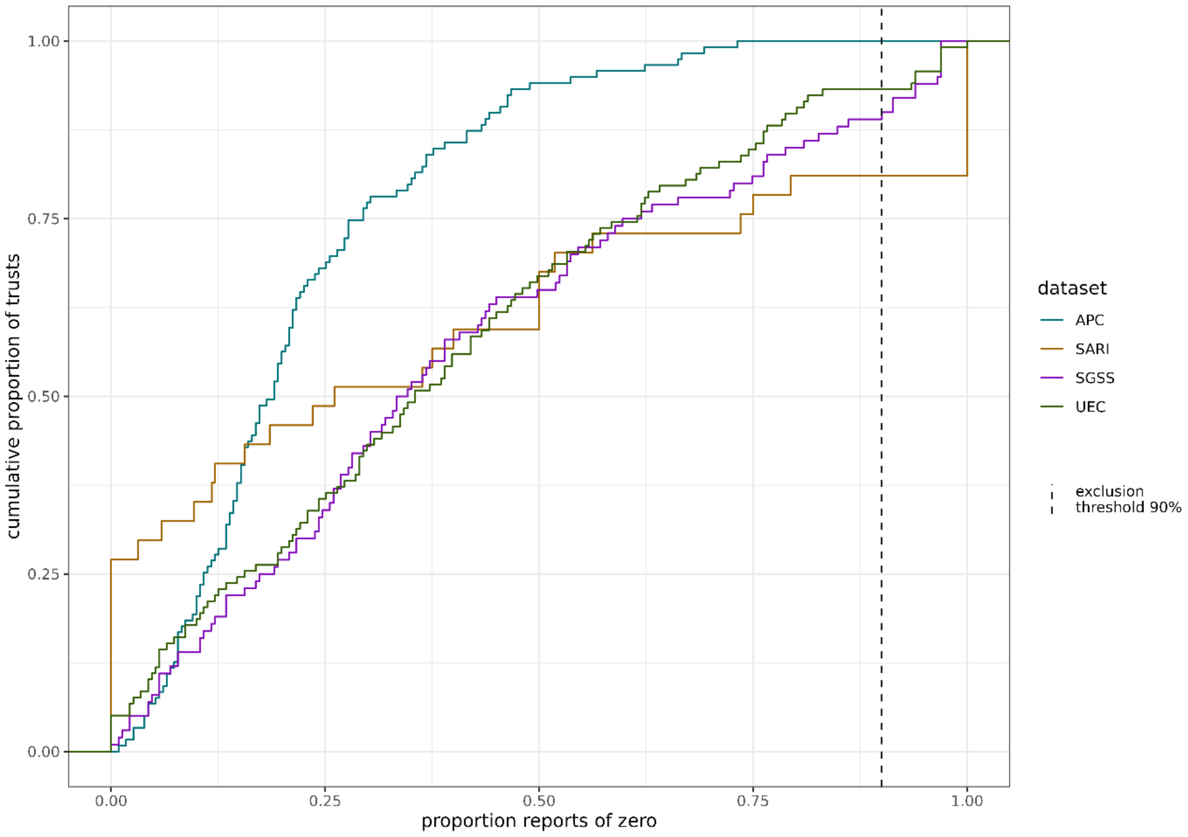


*Supplementary Figure 1. The proportion of trusts which have a set proportion of zeros reported over the study period. The weekly rolling mean value of the daily data (APC, SGSS, UEC) is shown to make it comparable to the weekly SARI. The cut-off threshold in the proportion of reports is used as an exclusion criteria for misreporting. A lower threshold would exclude more trusts that may have reported correctly across data sources, while a higher threshold would have limited impact.*

| Region | Dataset | Trusts reporting (minimum – maximum) | Peak Admissions | Cumulative Admissions |
|---|---|---|---|---|
| EAST OF ENGLAND | APC | 12 - 12 | 117 | 3371 |
| | SARI | 1 - 5 | 12 | 85 |
| | SGSS | 7 - 7 | 66 | 1216 |
| | UEC | 10 - 13 | 70 | 1829 |
| LONDON | APC | 18 - 18 | 223 | 6933 |
| | SARI | 2 - 4 | 81 | 483 |
| | SGSS | 12 - 12 | 60 | 1723 |
| | UEC | 10 - 17 | 60 | 2032 |
| MIDLANDS | APC | 20 - 20 | 311 | 8787 |
| | SARI | 0 - 2 | 29 | 151 |
| | SGSS | 16 - 16 | 306 | 8508 |
| | UEC | 15 - 18 | 244 | 6541 |
| NORTH EAST AND YORKSHIRE | APC | 21 - 21 | 292 | 10070 |
| | SARI | 1 - 3 | 41 | 229 |
| | SGSS | 13 - 13 | 224 | 7516 |
| | UEC | 15 - 19 | 251 | 8148 |
| NORTH WEST | APC | 16 - 16 | 230 | 6648 |
| | SARI | 3 - 4 | 23 | 106 |
| | SGSS | 13 - 13 | 186 | 3966 |
| | UEC | 11 - 13 | 139 | 4622 |
| SOUTH EAST | APC | 15 - 15 | 218 | 5724 |
| | SARI | 1 - 3 | 12 | 60 |
| | SGSS | 8 - 8 | 177 | 3277 |
| | UEC | 12 - 14 | 318 | 6505 |
| SOUTH WEST | APC | 13 - 13 | 206 | 4510 |
| | SARI | 1 - 3 | 37 | 122 |
| | SGSS | 9 - 9 | 145 | 3400 |
| | UEC | 10 - 12 | 133 | 3029 |

*Supplementary Table 1. Unmodelled NHS regional summaries of reported data across the different data sources. Counts are from the processed data after exclusion criteria are applied. Trust counts are the lowest and highest number of participating Trusts for a given report post exclusion criteria. Peak admissions are taken as the maximum admissions in each report and cumulative admissions the sum of all admissions reported. These metrics are not corrected for time varying participation and population catchment sizes.*

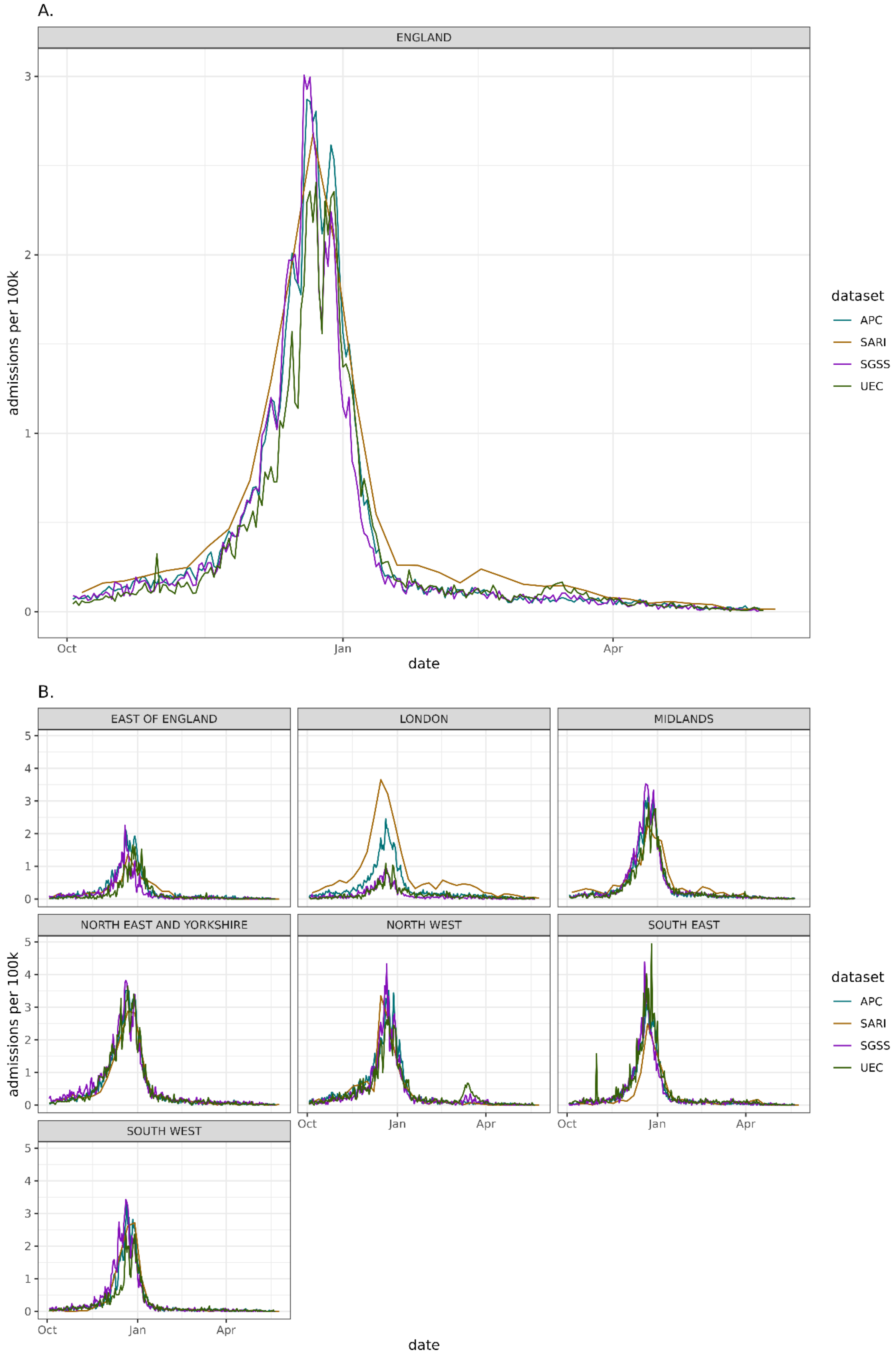


*Supplementary Figure 2. Influenza admission rate per 100k trust catchment population nationally (sub-plot A) and NHS commissioning region (sub-plot B) over the winter 2022/23 season.*

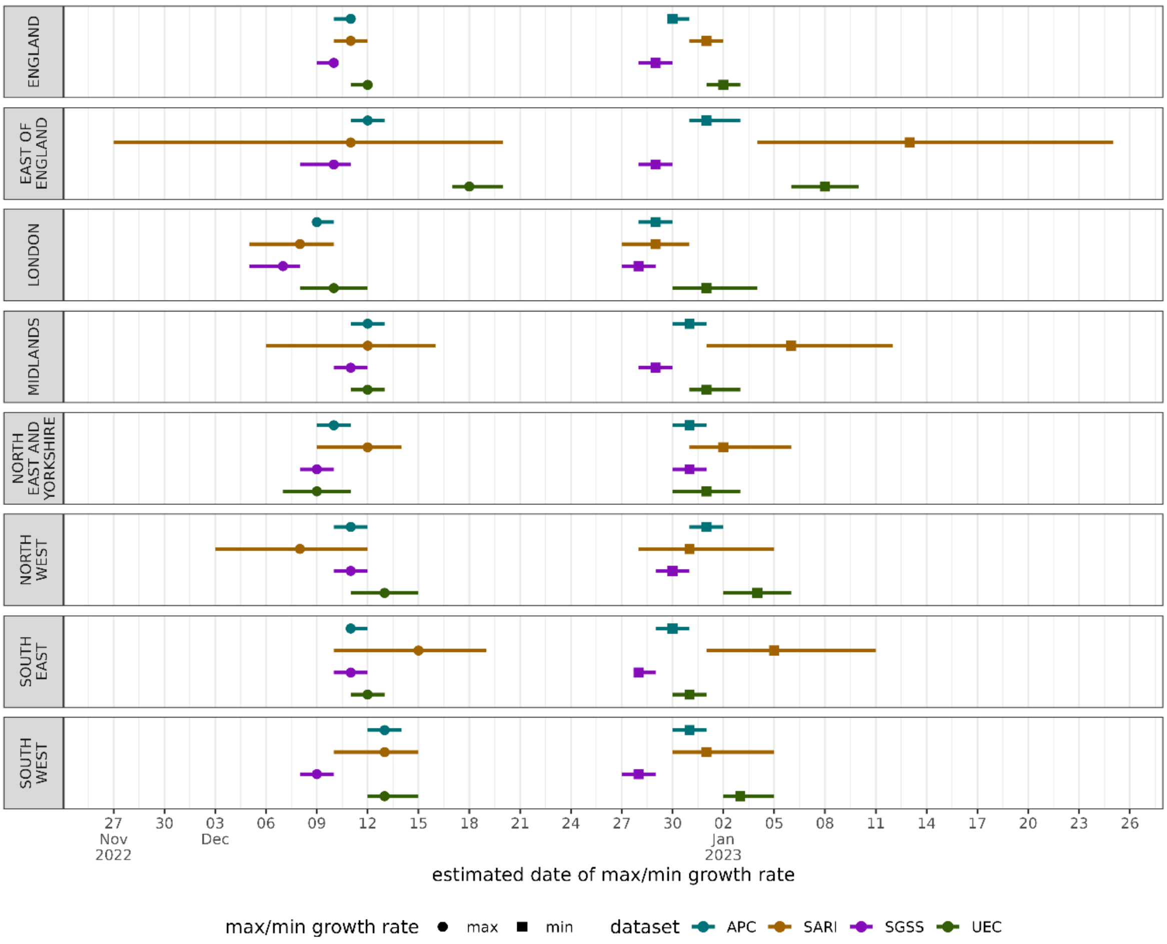


*Supplementary Figure 3. The estimated date of maximum and minimum growth rate, corresponding to the fastest increase and decreasing point of the epidemic wave across each data source over the 2022/23 winter season in England. The central point represents the median estimate, and the lines the 95% confidence interval.*

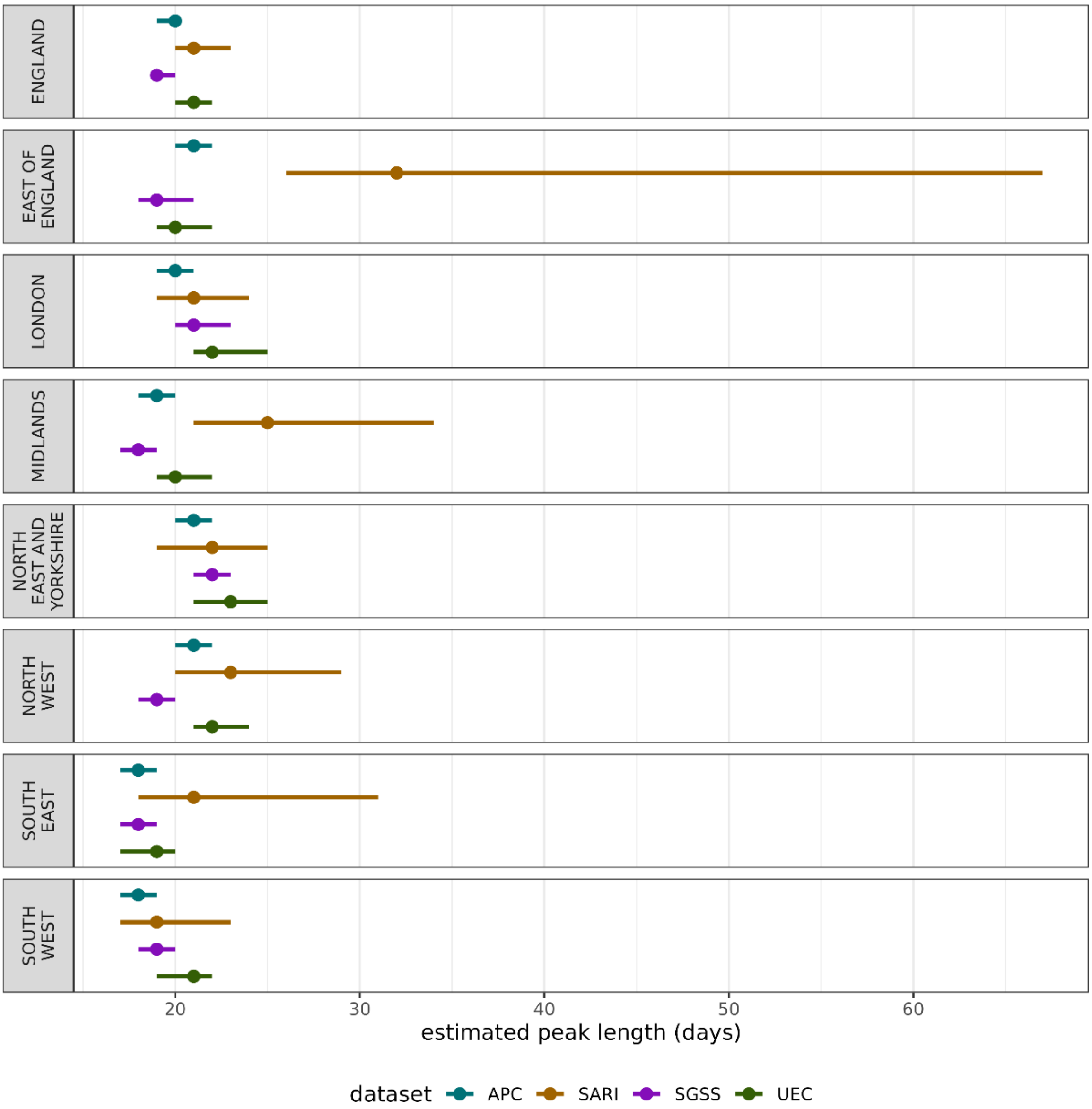


*Supplementary Figure 4. The estimated length of the peak, defined as the days between the date of maximum growth rate (fastest increase) and minimum growth rate (fastest decrease) across each data source over the 2022/23 winter season in England. The central point represents the median estimate, and the lines the 95% confidence interval.*
